# LARGE VOLUME SURFACTANT ADMINISTRATION IN PREMATURE INFANTS LESS THAN 35 WEEKS USING MINIMALLY INVASIVE SURFACTANT THERAPY

**DOI:** 10.1101/2022.08.20.22278507

**Authors:** Yahya Ethawi, Abrar Hussain, John Minski, Joe Miller, Ruben Alvaro

## Abstract

Large volume surfactant administration in premature infants less than 35 weeks using minimally invasive surfactant therapy (MIST).

**Background:** Surfactant in small volume preparations can be successfully administered to premature infants using a narrow-bore vascular catheter inserted into the trachea under direct vision. It was not known if surfactant in large volume preparations could be administered effectively to spontaneously breathing premature infants while maintaining them on nasal continuous positive airway pressure (nCPAP).

**Objective:** To evaluate the feasibility of administering a large volume of surfactant (5 ml/kg) in spontaneously breathing premature infants on nCPAP using a vascular catheter.

**Methods:** Single-arm interventional trial in two level III nurseries in Canada. Spontaneously breathing premature infants between 24 and 34 weeks’ gestation, born between July 4^th^, 2012 and September 5^th^, 2012 were eligible for the trial in the first 24 hours of life. A 16-gauge vascular catheter was inserted under direct vision through the vocal cords to administer 5 ml/kg of surfactant while maintaining the infants on nCPAP.

**Results:** Twenty-one premature infants were enrolled. The mean gestational age was 29 ± 3 (range 24 to 34 weeks) with mean birth weight of 1474 ± 575 g. Surfactant was successfully administered in 20 infants (95 %). There was a clear effect after surfactant administration with a decrease in oxygen requirement from 0.34 ± 0.111 before the procedure, to 0.26 ± 0.08 after the procedure (p = .001), and a decrease in nCPAP pressure from 7.2 ± 0.5 cm H_2_O before the procedure to 5.9 ± 0.5 after the procedure (p < .0001). The mean duration for surfactant administration was 8.3 ± 3.4 minutes. The procedure was temporarily stopped in three cases due to bradycardia and/or apnea with desaturations that spontaneously recovered without intervention. Coughing and reflux of the surfactant were transiently observed in four cases (19 %).

**Conclusion:** It is feasible to administer large volumes of surfactant without interrupting nCPAP via a vascular catheter, to premature infants less than 35 weeks’ gestation. Further trials are needed to confirm the safety and efficacy of this method compared to other methods of surfactant administration.

## Introduction

Exogenous surfactant administration is a recognized treatment for respiratory distress syndrome (RDS).^1^ For over 20 years it has been common practice to administer surfactant with positive pressure ventilation (PPV) to infants who manifest signs of surfactant deficiency.^2^ However, this approach has potential hazards associated with intubation: laryngospasm, bronchospasm, hemodynamic changes, raised intracranial pressure, and an increased risk of intracranial hemorrhage.^3^ Cchronic lung disease (CLD), defined as a need for oxygen at 36 weeks corrected gestation, remains a common poor outcome of premature infants.^4^ Although the etiology of CLD in premature infants is multifactorial,^4^ ventilator-induced lung injury remains one of the main implicated risk factors.^5^ Even a few PPV breaths may result in lung injury.^6^ A shift toward less invasive forms of respiratory support, like nCPAP, has been proposed in an effort to avert adverse pulmonary outcomes.^7^ Premedication drugs used prior to intubation for analgesia and/or muscle relaxation may affect the infant’s ability to sustain a proper respiratory drive, and may contribute to a delayed extubation once surfactant has been administered.^8^ Using non-invasive methods of respiratory support, and withholding or delaying surfactant administration to avoid intubation and mechanical ventilation, may also compromise the outcome.^9^

The intubation-surfactant-extubation (InSurE) procedure has been proposed by Victorin to preserve the well-documented benefits of surfactant therapy without the need for prolonged mechanical ventilation.^10, 11^ The InSurE procedure is attractive in principle, but complications associated with intubation and short-term PPV remains, especially in extremely premature infants. Although some studies suggested overall benefits of the InSurE procedure, ^12^ other studies failed to prove these benefits in that regard.^13^

In view of this, different methods of MIST have been introduced for evaluation: intra-amniotic instillation, ^14^ pharyngeal instillations, ^15^ administrations via laryngeal mask airway (LMA),^16^ and nebulized surfactant administration in spontaneously breathing infants.^17^

Other types of MIST using tracheal catheterization were described; the surfactant is given to spontaneously breathing premature infants via feeding tube^18, 19^ or vascular catheter^20^ placed in the trachea only during surfactant administration, and removed thereafter.

Kribs et al. described one of the MIST methods where they kept infants in need of surfactant on nCPAP and used a feeding tube to give the surfactant through the trachea.^18^ The technical difficulty associated with tracheal insertion of the flexible feeding tube, is that it mandates the use of Magill’s forceps to advance the tip of the feeding tube into the trachea. The use of additional instruments adds more difficulty and is a limitation to that procedure.

The research group at the University of Tasmania developed a technique of MIST (Hobart method) in which surfactant is administered into the trachea using a narrow-bore vascular catheter. This group has shown that a small volume of surfactant (1.25 ml/kg) can be effectively and safely administered in premature infants using this technique.^20^ A less concentrated, larger volume of surfactant, bovine lethicin extracted surfactant (BLES), in a dose of 5 ml/kg is available in different neonatal units, including Canada’s,and Survanta in a dose of 4 ml/kg in many other countries. It is not known whether this technique can be applied effectively using BLES.

The purpose of this pilot study was to evaluate the feasibility of administrating 5 ml/kg of a less concentrated, larger volume of surfactant, BLES, in spontaneously breathing premature infants using a 16-gauge vascular catheter, while maintaining them on nCPAP. We hypothesized that early CPAP and large volume minimally invasive surfactant (ECALMIST) administration using a narrow-bore vascular catheter was feasible, safe, and effective.

## Methods

### Study Design

This was a prospective non-randomized feasibility clinical trial conducted at two centers (Health Sciences Centre, and the St. Boniface General Hospital) in Winnipeg, Manitoba, Canada. The study was approved by the research ethics committees in the two centers and by the faculty committee for the use of human subjects in research. The study was registered at http://www.clinicaltrial.gov, registration number NCT01553292. The study was conducted over a four month period from July 4^th^, 2012 to September 5^th^, 2012, and included premature infants between 24 and 34 weeks gestation, within the first 24 hrs of life, and with a clinical diagnosis of respiratory distress thought to be due to surfactant deficiency; the clinical decision to give surfactant was made by the treating team. The participants had to demonstrate that they were able to breathe spontaneously on nCPAP. An informed written consent was obtained from at least one of the parents prior to enrollment. Infants with major congenital anomalies or who required mechanical ventilation (not spontaneously breathing) were excluded.

### Procedure

The type of catheter used was a 16 gauge (5 French) vascular catheter (Angiocath, BD, Sandy, Utah, USA). It was prepared by marking a point with tape indicating the desired depth of insertion at the level of the lips (6 cm plus weight of the infant in kg). BLES (5 ml/kg) was drawn up in a syringe and warmed to body temperature. The vascular catheter was inserted using direct laryngoscopy and an appropriate size Miller laryngoscope blade through the vocal cords under direct vision. When available, the laryngoscope was connected to a video camera to record the procedure. After catheter placement, the laryngoscope blade was removed and the catheter stabilized by two fingers at the level of the upper lip. The surfactant syringe was connected to the catheter hub either before or after inserting the vascular catheter into the trachea according to the preference of the person who was performing the procedure. Once the vascular catheter was correctly positioned, a small volume of surfactant (0.25 - 0.5 ml) was administered. The syringe was then disconnected from the vascular catheter to observe the fluctuation of BLES up and down with respiration as indication of accurate intubation of the trachea. Once placement was confirmed and the initial volume was absorbed, the remaining volume of BLES was slowly administered in small aliquots, (0.25-0.5 ml at a time over 20-30 seconds) with approximately 10 seconds between each aliquot. At the end of the procedure, the operator flushed the vascular catheter with 0.5 ml of air before removing it. In all infants, an orogastric tube was inserted after the procedure, and the stomach was aspirated to ensure that surfactant was not accidentally given into the stomach. During the procedure, if the infant went apneic, bradycardic, or developed a significant decrease in oxygen saturation (< 70 %), the administration of BLES was stopped for 20 to 30 seconds until the vital signs returned to baseline, then the procedure was resumed. If the infant could not recover, and required other resuscitative measures, inspired oxygen (FiO_2_) and nCPAP pressure were increased and/or PPV was provided using bag and mask ventilation as per the discretion of the attending physician. If vital signs remained unstable, or the clinical condition further deteriorated, the procedure was terminated and the infant was intubated and ventilated as per the neonatal resuscitation program (NRP) guidelines.

### Data Collection

Demographic and clinical data were collected prospectively for all infants. The number and duration of all tracheal catheterization attempts, as well as the total time of the procedure, was collected. Vital signs during the procedure were recorded; these included heart rate (HR), respiratory rate, and oxygen saturation (recorded from Masimo Radical 7 pulse oximeter). Changes in FiO_2_ requirement and changes in nCPAP pressure for the first four hours after the procedure were also documented and collected. The need for mechanical ventilation was reported at 24 and 72 hours. Data from the Masimo monitor was transferred to a computer and analyzed using the PROFOX computer program (PROFOX Associates, Inc. 651 E. Pennsylvania Ave, Suite 101 Escondido, CA 92025) figure5. The procedure was videotaped when permitted, after parental consent. An index, arbitrarily named *CPAP index*, was calculated before and after the procedure. The purpose of this index was to show the changes of the three parameters together: The FiO_2_ in decimal, the CPAP pressure in centimeters of water, and the oxygen saturation in decimal as measured by the Masimo radical 7 pulse oximeter. The CPAP index was calculated using the following formula: FiO_2_*CPAP/SpO_2_*100. This formula is the same formula used for calculation of Oxygen Index (OI) which is FiO_2_*mean airway pressure (MAP) / Blood oxygen level (PO_2_)

1. The MAP that is used in the OI formula, which is measured by ventilator during mechanical ventilation, was replacing by CPAP pressure measured in cm of water in the CPAP index.
2. The PO_2_ measured by blood gas analysis in the OI formula, was replaced by oxygen saturation measured by the Masimo pulse oximeter in the CPAP index.
3. To make the resulted number shorter, the result was divided by 100.

All adverse events of the procedure were collected, including reflux of the surfactant, apnea, bradycardia, and decrease in oxygen saturation. Also, the need to temporarily stop or to terminate the procedure, increase the FiO_2_ requirement, changes in nCPAP pressure, and any other not listed adverse event that arose during the procedure, were collected. Significant bradycardia was defined as HR < 100 per minute for more than 5 seconds, HR < 100 per minute associated with respiratory compromise that required PPV, or HR < 60 per minute. Significant decrease in oxygen saturation was defined as any oxygen saturation below 75 %, or below 85 % for more than 20 seconds. Apnea was defined as a respiratory pause for ≥ 20 seconds, or any respiratory pause associated with a decrease in oxygen saturation below 85 or HR below 100 beats per minute. Failure of tracheal catheterization was defined as two unsuccessful attempts after 20 seconds for each attempt.

The percentage, mean, median, and range calculations were used to describe the demographic data. Two-sample t-test for paired samples, and z-test with confidence interval of 95 % and alpha level of 0.5 were used to measure the significant difference between CPAP pressure, FiO_2_ requirement, saturation, and CPAP index before and after the procedure.

## Results

During the study period, 21 premature infants of gestational age between 24 weeks 0 days and 34 weeks 6 days with respiratory distress at birth were enrolled out of 41 less than 25 weeks’ premature infants after an informed written consent was obtained from at least one of the parents. Out of 41 premature infants born during the study period, 21 were recruited. The remaining 20 were excluded due to congenital malformations in 2 infants, the need for PPV in 3 infants, and the need for mechanical ventilation in 4 infants. The parents of the remaining 11 infants declined to consent (figure 1, table 3). The characteristics of recruited infants are showed in table 1, while table 2 shows the characteristics of the ECALMIST procedure regarding place, personnel, number of trials, complications, and duration of the procedure.

**Table 1:** Characteristics of the newborns enrolled in the study.

|  |  |
| --- | --- |
| Birth weight – g±2SD | 1419 ± 482 |
| Gestational age - wk±2SD (Range) | 29 ± 2 (24-34+6) |
| Female sex – no. (%) | 18 (51) |
| Male sex – no. (%) | 9 (49) |
| Apgar score at 5 min – median (range) | 8 (5-10) |
| Cesarean section – no. (%) | 19 (54) |
| Antenatal steroid –no. (%) | 23 (68) |

**Table 2:** characteristics of the procedure.

|  |  |
| --- | --- |
| Postnatal Age - minutes±1SD | 35 ± 17 |
| Delivery room – no. (%) | 12 (34) |
| NICU – no. (%) | 23 (66) |
| Neonatal fellow - no. (%) | 21 (60) |
| Senior house medical officer - no. (%) | 7 (20) |
| Respiratory therapist - no. (%) | 5 (14) |
| Neonatologist - no. (%) | 2 (6) |
| Duration – minutes (range) | 6.9 ± 3.1 (2 – 16) |
| First attempt - no. (%) | 27 (77) |
| Second attempt - no. (%) | 7 (20) |
| Failed catheterization - no. (%) | 1 (3) |
| Bradycardia – no. (%) | 2 (6) |
| Desaturation – no. (%) | 3 (8.6) |
| Reflux of surfactant – no. (%) | 7 (20) |

**Table 3:**
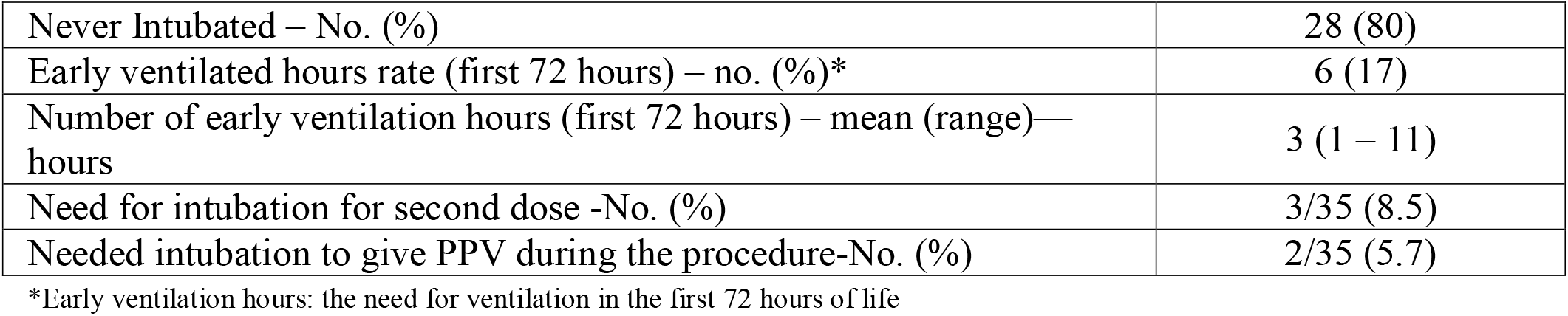
Ventilation outcomes.

|  |  |
| --- | --- |
| Never Intubated – No. (%) | 28 (80) |
| Early ventilated hours rate (first 72 hours) – no. (%)* | 6 (17) |
| Number of early ventilation hours (first 72 hours) – mean (range)—<br>hours | 3 (1 – 11) |
| Need for intubation for second dose -No. (%) | 3/35 (8.5) |
| Needed intubation to give PPV during the procedure-No. (%) | 2/35 (5.7) |

\*Early ventilation hours: the need for ventilation in the first 72 hours of life

**Figure 1:**
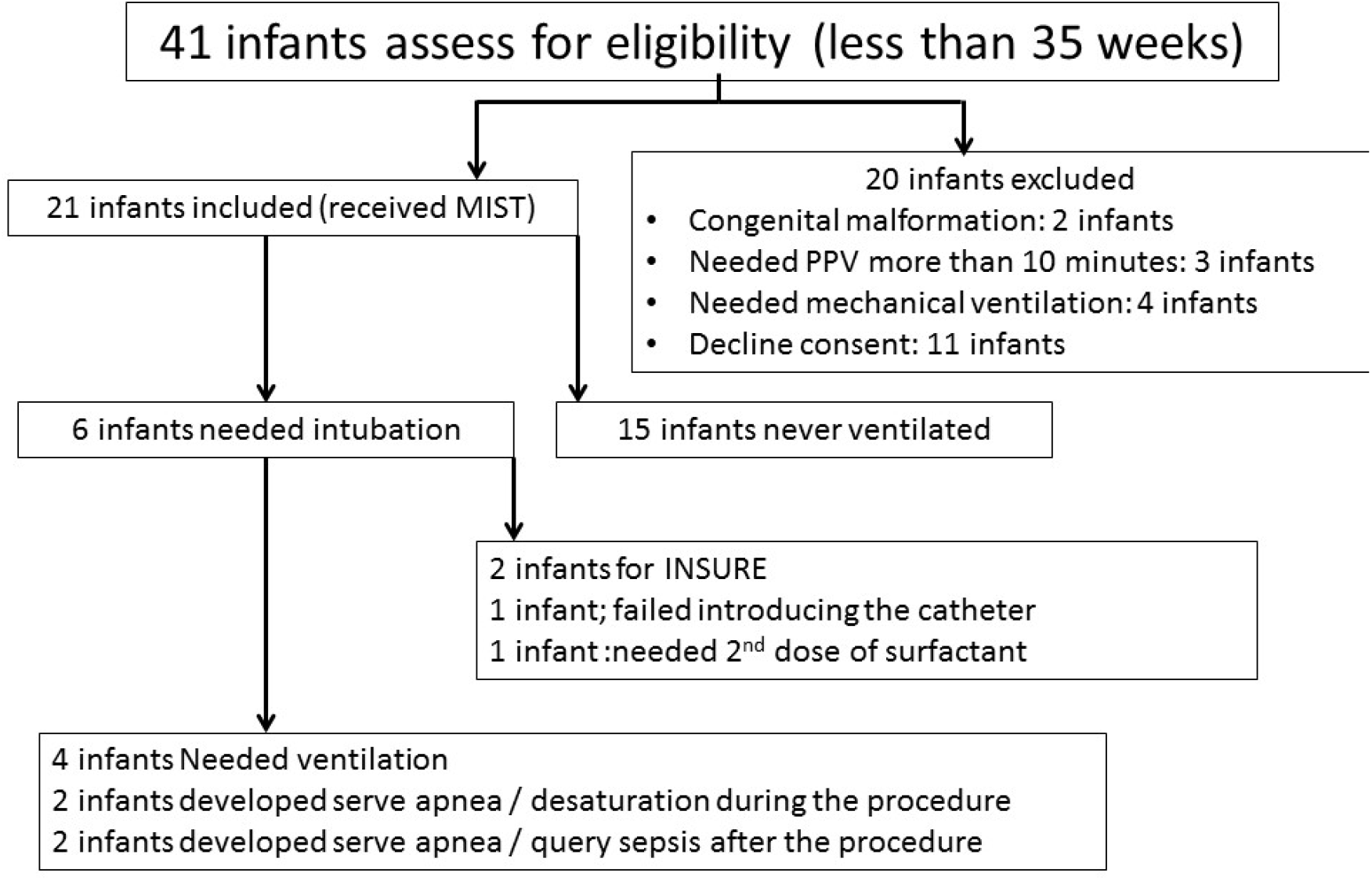
Trial profile

The effect of surfactant administration was observed with the ECALMIST procedure. The nCPAP pressure (figure 2, figure 4), the oxygen requirement (figure 3, figure 4), and the CPAP index (figure 4), were reduced after the procedure in comparison to prior to the procedure (P < .0001, P = .004, P < .00001 respectively). During the procedure, a significant decrease in heart rate (HR < 100 beats/min) was observed in only 1 out of 21 patients (4.8 %), and a decrease in oxygen saturation below 70 was observed in 2 out of 21 patients (9.5 %).

**Figure 2:**
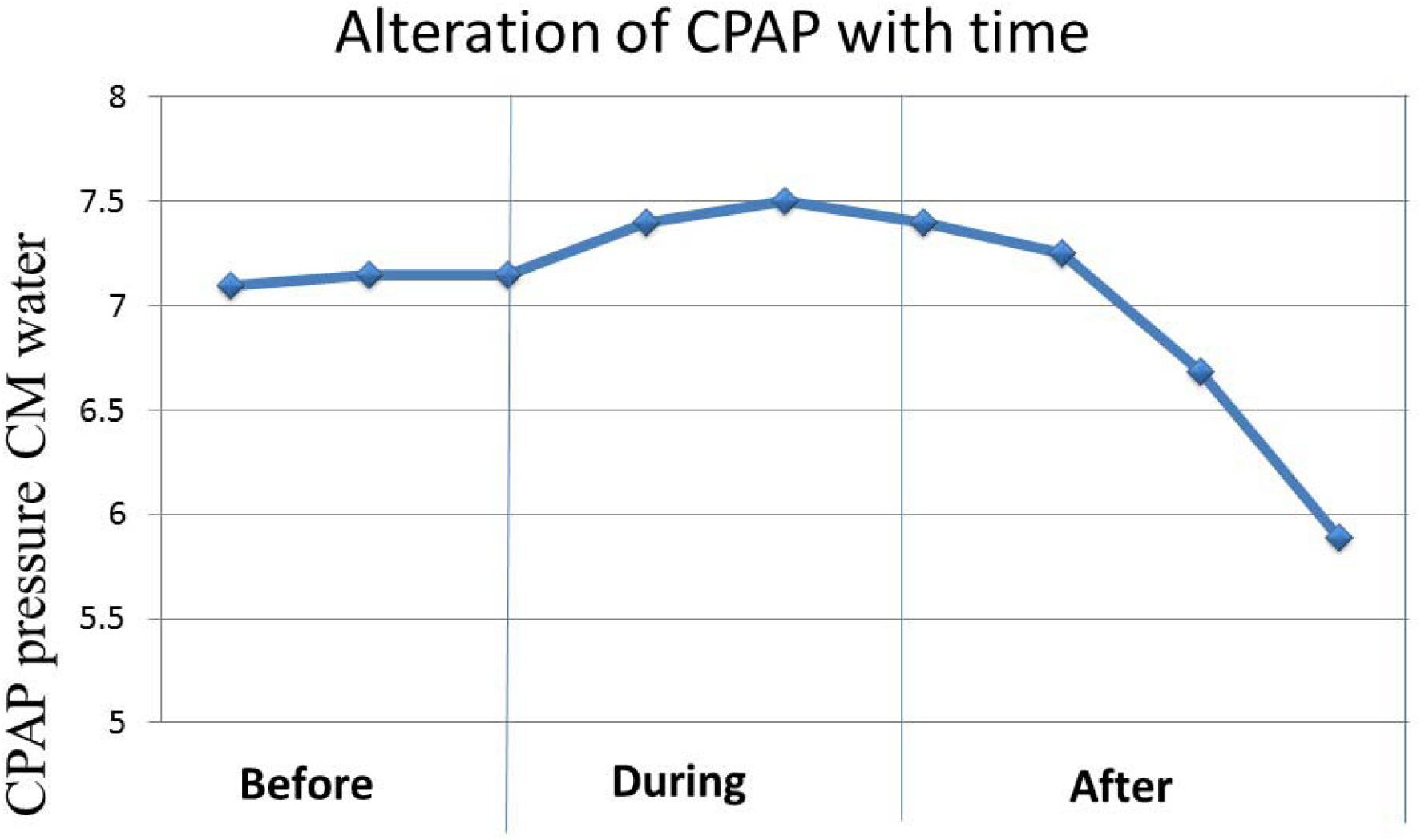
FIO_2_ significantly decreases immediately after the administration of surfactant through the vascular catheter

**Figure 3:**
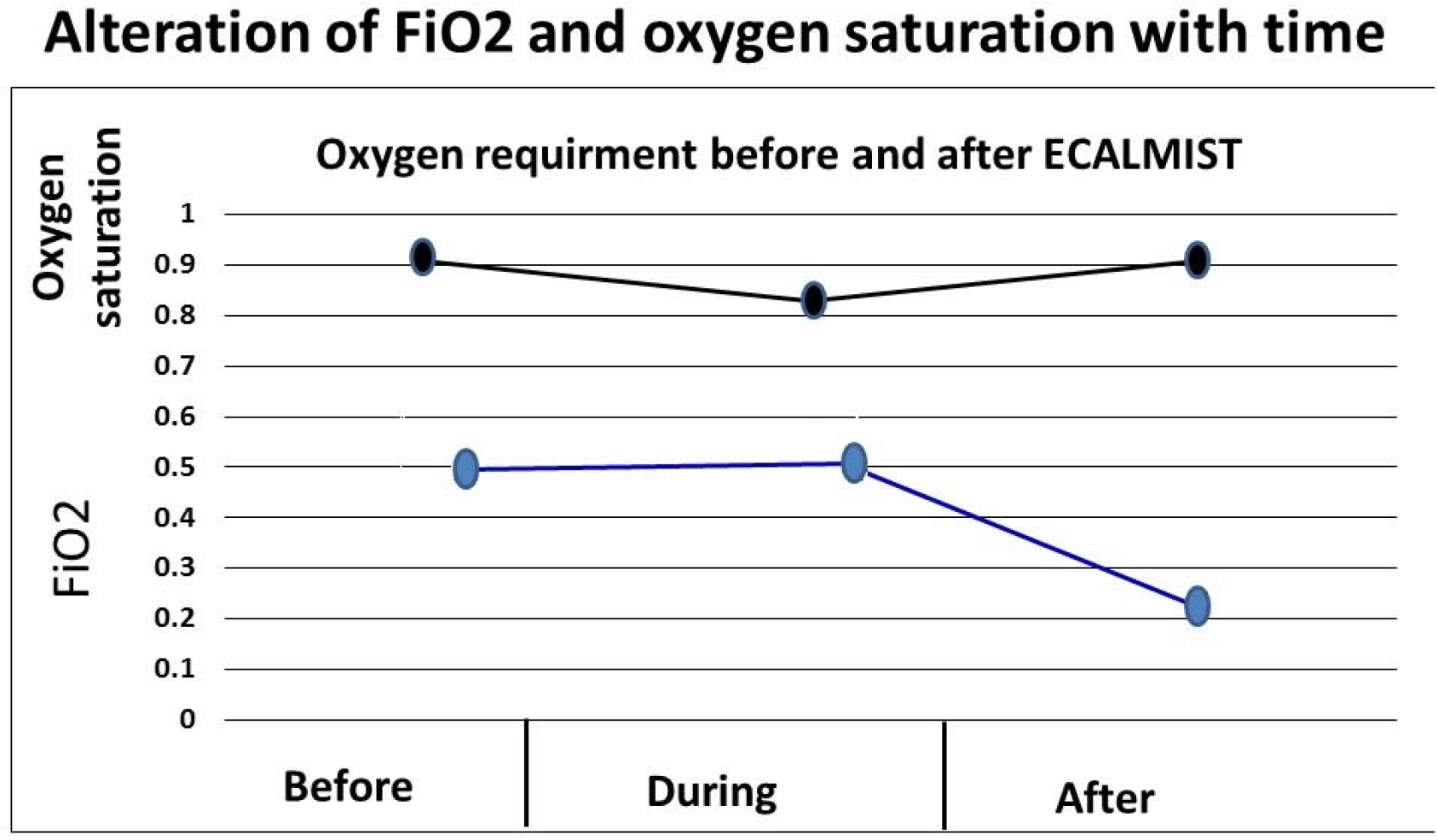
CPAP pressure significantly decreases within 4 hours after the administration of surfactant through the vascular catheter.

**Figure 4.**
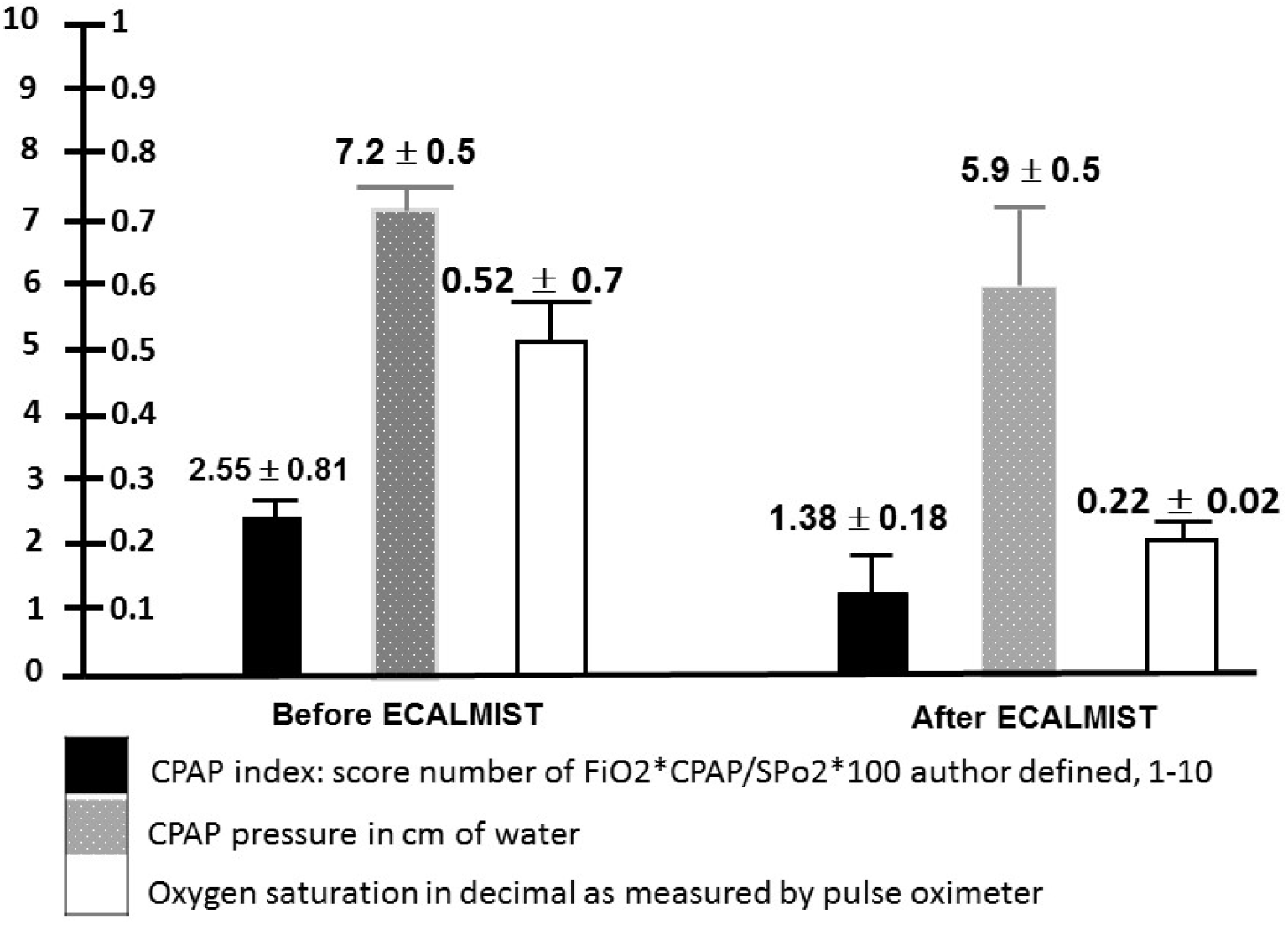
nCPAP pressures, CPAP index and oxygen requirement changes before and after the procedure

**Figure 5:**
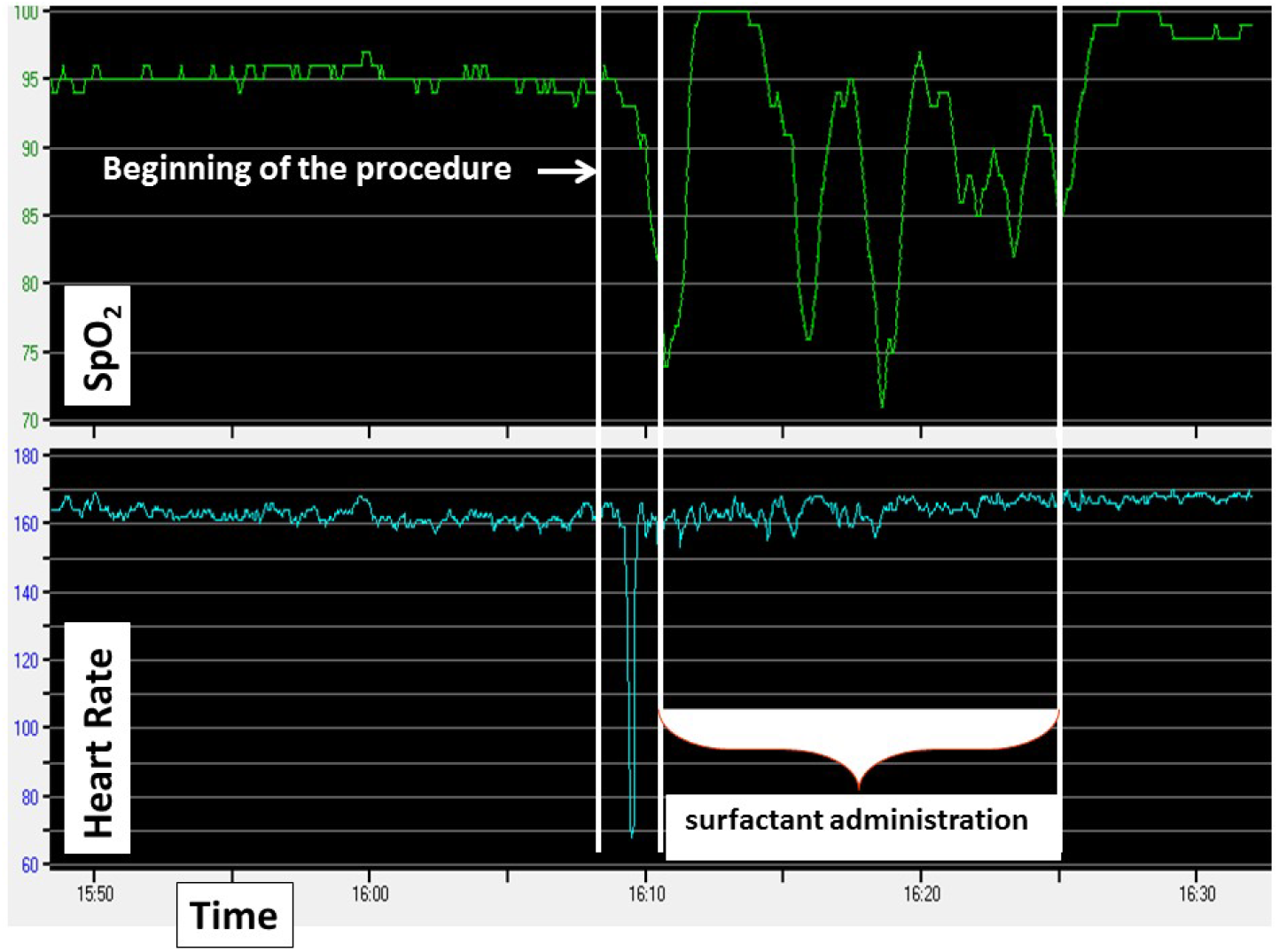
Representative Tracing: A brief and very transient bradycardia developed during the introduction of the vascular catheter that resolved spontaneously as soon as the laryngoscope was removed. Fluctuations in oxygen saturation were observed during the administration of surfactant that resolved as soon as the procedure ended. The heart rate remained stable throughout.

Other associated neonatal outcomes in these patients occurred: patent ductus arteriosus (PDA) in 2 out of 21 patients (9.5 %), intraventricular hemorrhage (IVH) grade I in 2 out of 21 patients, IVH grade II in 1 out of 21 patients, periventricular leukomalacia (PVL) in 1 out of 21 patients, pneumothorax in 1 out of 21 patients, and retinopathy of prematurity (ROP) in 3 out of 21 patients (14.28 %), as shown in table 4.

**Table 4:** Associated neonatal outcome.

|  |  |
| --- | --- |
| PDA – No. (%) | 4 (11.4) |
| NEC (necrotizing enterocolitis) – No. (%) | 1 (2.9) |
| IVH (Intraventricular hemorrhage) grade I-No.(%) | 4 (11.4) |
| IVH (Intraventricular hemorrhage) grade II-No.(%) | 2 (5.7) |
| IVH (Intraventricular hemorrhage) grade I-No.(%) | 1 (2.9) |
| PVL (periventricular leukomalacia-No (%) | 2 (5.7) |
| Pneumothorax | 1(2.9) |
| ROP (Retinopathy of prematurity)-No.(%) | 5 (14.3) |
| Death | 1(2.9) |
| CLD (need for FiO2 at 36 weeks corrected<br>gestational age0-No. (%) | 6 (17.14) |
| Composite outcome (death and CLD) | 7(20%) |

## Discussion

In this pilot study, we evaluated the feasibility of administering a large volume of surfactant (5 ml/kg) in spontaneously breathing premature infants between 24 to 35 weeks gestation on nCPAP using a 17 gauge vascular catheter. The ECALMIST method was successful in 20 out of 21 infants enrolled in the study while maintaining the nCPAP. Previous studies of MIST methods used small volume concentrated surfactant (1.25 - 3 ml/kg).^17,18,20^ In the current study, we demonstrated that administration of a large volume of surfactant is feasible using the ECALMIST method. There was evidence of the effects of surfactant delivery to the alveoli, with an immediate physiological benefit including rapid and sustained reduction in oxygen requirement and nCPAP pressure. The procedure was not successful in 1 out of 21 infants; this trial was attempted by a newly trained respiratory therapist. After this first attempt, the infant was not stable enough to try another trial by a more experience operator, so the managing team decided that the infant should be intubated to administer surfactant. These findings support our hypothesis that it is feasible to use MIST using a narrow-bore vascular catheter to administer a large volume (5 ml/kg) of surfactant while the infant is maintained on nCPAP (ECALMIST).

This study is the first to evaluate the feasibility of administering large volume (5 ml/kg) surfactant combined with the use of nCPAP through a small catheter in premature infants with respiratory distress due to surfactant deficiency. As judged by the high proportion of cases requiring only a single catheterization attempt (86 %), the technique used in the present study was simple and easy to perform by health professionals reasonably trained in neonatal intubation. The failure of intubation and the need for a second trial mainly occurred with health care providers who had less than 2 years of experience with neonatal intubation. When compared to the standard intubation with an endotracheal tube, the narrow bore and semi-rigid design of the catheter allows for an easy pass through the vocal cords without obstructing the view of the glottis. The longer duration of the ECALMIST procedure compared to other MIST studies that used small volumes, was likely attributed to the larger volume of surfactant used in the present study. The episode of bradycardia observed during the procedure was not different from those observed during standard neonatal intubation without premedication occurring during the manipulation of the laryngoscope blade to obtain a good view of the vocal cords. By reviewing videos of delivery room resuscitation, O’Donnell et al. observed that standard intubation attempts in infants are often unsuccessful (up to 38 %) compared to 14 % in our study, and successful attempts frequently require > 30 seconds compared to < 30 seconds in our study. These authors also showed that during these standard intubation procedures, 49 % of infants deteriorated with a decrease in oxygen saturation by ≥ 10 %, while in our study it was 28.6 % (only in 9.5 % of these cases was the oxygen saturation below 70 %). They also showed in 33 % of infants, a decrease in HRby ≥ 10 % in 15 % of the cases, and a decrease in both oxygen saturation and HRby ≥ 10 % in 44 % of the cases. In our study, a significant decreased in HR (< 100 beats per minute for more than 5 seconds) was observed in only 1 patient out of 21 (4.8 %). A decrease in both oxygen saturation ≥ 10 % and HRwas observed in 28.6 % of cases during the procedure, while a decrease in both HR below 100 beats per minute and oxygen saturation below 70 % was observed in 14 % of the cases. The first successful trial of catheterization of the trachea in our study was observed in intubation in our study in 86 %, while Simon et al., in a prospective study of 75 neonatal and pediatric intensive care units, documented that the rate of successful conventional neonatal intubation was only 58 %. These authors showed that neonatal intubation attempts were associated with an incidence of 31 % desaturations, and 24 % desaturations and bradycardias.

Kribs et al. in a retrospective, multicenter data, suggests that the application of surfactant to spontaneously breathing infants is associated with low rates of mechanical ventilation, bronchopulmonary dysplasia (BPD), and the combined outcomes of death or BPD at 36 weeks postmenstrual age. Based on several recent studies, it is now evident that initial stabilization with nCPAP and rescue surfactant treatment is not worse than intubation, mechanical ventilation, and prophylactic surfactant administration right after birth. However, a combination of nCPAP with very early surfactant administration through a minimally invasive technique, MIST, can lead to better clinical outcomes by avoiding intubation and exposure to mechanical ventilation.

Different methods of MIST using small volume concentrated surfactant is used in some NICUs with the objective of avoiding intubation and decreasing the exposure of premature infants to the negative effects of intubation and mechanical ventilation. Kribs et al. showed that the administration of surfactant via a thin small gastric tube to spontaneously breathing premature infants receiving CPAP, reduced the need for mechanical ventilation. In this study, the thin catheter was placed in the trachea with use of Magill forceps with direct visualization of the vocal cords with a laryngoscope. The surfactant was then administered during a 1 to 3-minute time span. Dargaville et al.^20^ found that a small volume concentrated surfactant (1.25 ml/kg) could be successfully administered to premature infants without premedication using a narrow-bore vascular catheter passed into the trachea under direct vision without using Magill forceps. Using this technique, these authors found a clear therapeutic benefit with reductions in FiO_2_ and nCPAP pressure. With modification of MIST by vascular catheter to adapt large volume surfactant in our procedure, checking the accuracy of catheter placement by observing the surfactant moving up and down after disconnecting the syringe from the catheter, giving the surfactant on boluses, maintaining the vital signs, and flushing the catheter with 0.5 ml of air at the end of the procedure, MIST can be used to deliver large volume surfactant. Although our present study has the limitation of being non-randomized and involves a relatively small number of premature infants, the ECALMIST technique could be an alternative approach to administering a large volume of surfactant without the need for intubation and PPV.

## Conclusion

In premature infants < 35 weeks’ gestation with RDS, a volume of 5 ml/Kg of surfactant (BLES) can be delivered via a small vascular catheter without interrupting nCPAP. The technique can be done by health professionals trained in neonatal intubation. Administration of surfactant using the ECALMIST technique may reduce the need for mechanical ventilation in very low birth weight infants, potentially decreasing the risk of CLD. Further randomized controlled studies are needed to compare the short and long-term outcomes of ECALMIST with the standard approaches to premature infants with surfactant deficiency at birth.

## Data Availability

All data produced in the present study are available upon reasonable request to the authors

## Acknowledgment

The study group would like to thank the families for their willingness to enroll their babies in this study. Special thanks also goes out to our colleagues for their help in finishing this trial.

